# Adult Life Course Trajectories of Lung Function and the Development of Interstitial Lung Abnormalities: The CARDIA Lung Study

**DOI:** 10.64898/2026.03.03.26347486

**Authors:** Kevin M. Grudzinski, Gabrielle Y. Liu, Laura A. Colangelo, Kavitha C. Selvan, Rachel Putman, Gary M. Hunninghake, Raul San Jose Estepar, George Washko, Ravi Kalhan, Anthony J. Esposito

## Abstract

**Background:** Interstitial lung abnormalities (ILA) are radiologic findings of increased lung density or fibrosis in individuals without clinical interstitial lung disease (ILD) and are associated with increased mortality and progression to ILD. Understanding physiologic trajectories of lung function preceding ILA diagnosis may illuminate early mechanisms of lung injury.

**Methods:** We recruited participants from the Coronary Artery Risk Development in Young Adults (CARDIA) Lung Study, a prospective cohort of adults enrolled at ages 18–30 years and followed longitudinally for 25 years. Percent predicted forced vital capacity (ppFVC) was measured at five study visits over 20 years. Individual ppFVC trajectories were estimated using random coefficient models. Person-specific slopes were incorporated into logistic regression models to examine associations with visually detected ILA on chest CT at exam year 25. Models were adjusted for age, sex, race, body mass index, pack-years of smoking, and study center.

**Results:** Among 3,136 participants with complete data, 57 (1.8%) had ILA at mean age 51 years. In univariable and multivariable models, individuals with ILA had greater cumulative decline in ppFVC over the 20 years preceding diagnosis. Each 10% absolute decline in ppFVC was associated with more than twice the odds of ILA (adjusted OR 2.21, 95% confidence interval 1.47–3.31, p = 0.0001).

**Conclusions:** Greater longitudinal decline in FVC from early adulthood was strongly associated with the presence of ILA at midlife. These findings suggest that physiologic impairments precede radiologic evidence of subclinical parenchymal lung abnormalities, underscoring the potential of life course lung function trajectories to identify individuals at risk for developing ILD.

## BACKGROUND

Interstitial lung abnormalities (ILA) are patterns of increased lung density and/or fibrosis on chest computed tomography (CT) and are associated with respiratory symptoms, progression to interstitial lung disease (ILD), and increased mortality.^1–5^ Typically, lung function peaks in early adulthood and declines with age. Trajectories of lung function decline may vary by the timing and amplitude of an individual’s peak and subsequent rate of decline. Those with low peak and/or rapid rate of decline in lung function are at increased risk of developing chronic obstructive pulmonary disease and non-respiratory conditions.^6–9^ Interrupting the development of ILD necessitates identifying those at highest risk. Previous studies, including our group’s work in the Coronary Artery Risk Development in Young Adults (CARDIA) cohort, has demonstrated that impaired early-adult forced vital capacity (FVC) trajectories are associated with increased risk of quantitative interstitial abnormalities (QIA) on CT.^9–10^ While QIA provide valuable, machine learning–derived measures of diffuse lung density changes, they are more prevalent and may reflect broader subclinical or nonspecific findings. In contrast, visually detected ILA align with consensus definitions of early ILD and are more directly linked to clinical outcomes such as disease progression and mortality, offering complementary but greater clinical relevance for hypothesis generation. We hypothesized that greater lifetime decline in FVC is associated with increased risk of visually detected ILA. Understanding factors that predispose individuals to poor lung health preceding diagnosis of ILA may generate hypotheses about early mechanisms involved in ILD pathogenesis.

## METHODS

The CARDIA Lung Study is a longitudinal cohort study that commenced in 1985 with 5,115 adults from four communities in the United States. CARDIA is reviewed annually by institutional review boards at each center. Participants were aged 18-30 years at baseline and were recruited equally between self-reported White and Black race.^11^ Lung function was assessed using race-neutral reference equations at five visits over twenty years.

CT examinations of the chest obtained at exam year 25 (Y25) were visually assessed for ILA per the American Thoracic Society recommendations utilizing a three-reader method.^12–13^ ILA was categorized as none, indeterminate, and definite. Indeterminate ILA cases were excluded from analyses.

Linear random coefficient models (RCM) were applied to model trajectories of percent predicted FVC (ppFVC) as a function of age. FVC was measured at various timepoints at exam year zero through twenty. Y25 and subsequent exam years were excluded as ILA was assessed on CT at Y25. Person-specific slopes from the RCMs were used as independent variables in univariable and multivariable logistic regression models assessing associations of longitudinal ppFVC trajectories with the outcome of Y25 ILA. Covariates included age, race, sex, body mass index (BMI), pack-years of smoking, and center. These covariates were assessed at Y25 for all analyses.

## RESULTS

Of 5,115 baseline CARDIA participants, 3,136 were included in analyses (**Figure 1**). ILA prevalence at Y25 was 1.8% (N=57). Of those with ILA, 27 participants (47.4%) were male, and 19 (33.3%) were White race. Ever (current or former) tobacco smokers varied between 33.3% in the no ILA group and 59.6% in the ILA group. The characteristics of those with no ILA and ILA at Y25 are displayed in **Table 1**. Greater life-course decline in FVC was associated with higher odds of Y25 ILA with an adjusted OR 2.21 (95% confidence interval 1.47-3.31, p=0.0001) per decline in ppFVC of 10% over the preceding twenty-year observation period (**Figure 2A**).

**Table 1.** Demographic and clinical characteristics of the study participants with no ILA and definite ILA at CARDIA exam year 25.

|  | No ILA | Definite ILA |
| --- | --- | --- |
| **N (%)** | 2807 (98.0) | 57 (2.0) |
| **Age, mean (SD)** | 50.0 (3.6) | 51.3 (3.3) |
| **Sex, male, N (%)** | 1196 (42.6) | 27 (47.4) |
| **Race, White, N (%)** | 1509 (53.8) | 19 (33.3) |
| **BMI kg/m<sup>2</sup>, mean (SD)** | 30.1 (7.1) | 30.9 (7.9) |
| **Lifetime pack-years of cigarette smoking, median [Q1, Q3]** | 0 [0, 6.3] | 8.6 [0, 23.0] |
| **Smoking Status (Y20)** |  |  |
| Never, N (%) | 1592 (63.0) | 14 (29.2) |
| Former, N (%) | 501 (19.8) | 8 (16.7) |
| Current, N (%) | 434 (17.2) | 26 (54.2) |
| **Baseline ppFVC (%)** | 100.9 | 98.7 |
| **Y20 ppFVC (%)** | 96.2 | 86.9 |

Abbreviations: BMI = body mass index; ILA = interstitial lung abnormalities; Q1 = 1<sup>st</sup> quartile; Q3 = 3<sup>rd</sup> quartile; SD = standard deviation; Y20 = CARDIA exam year 20; ppFVC = percent predicted FVC.

**Figure 1.**
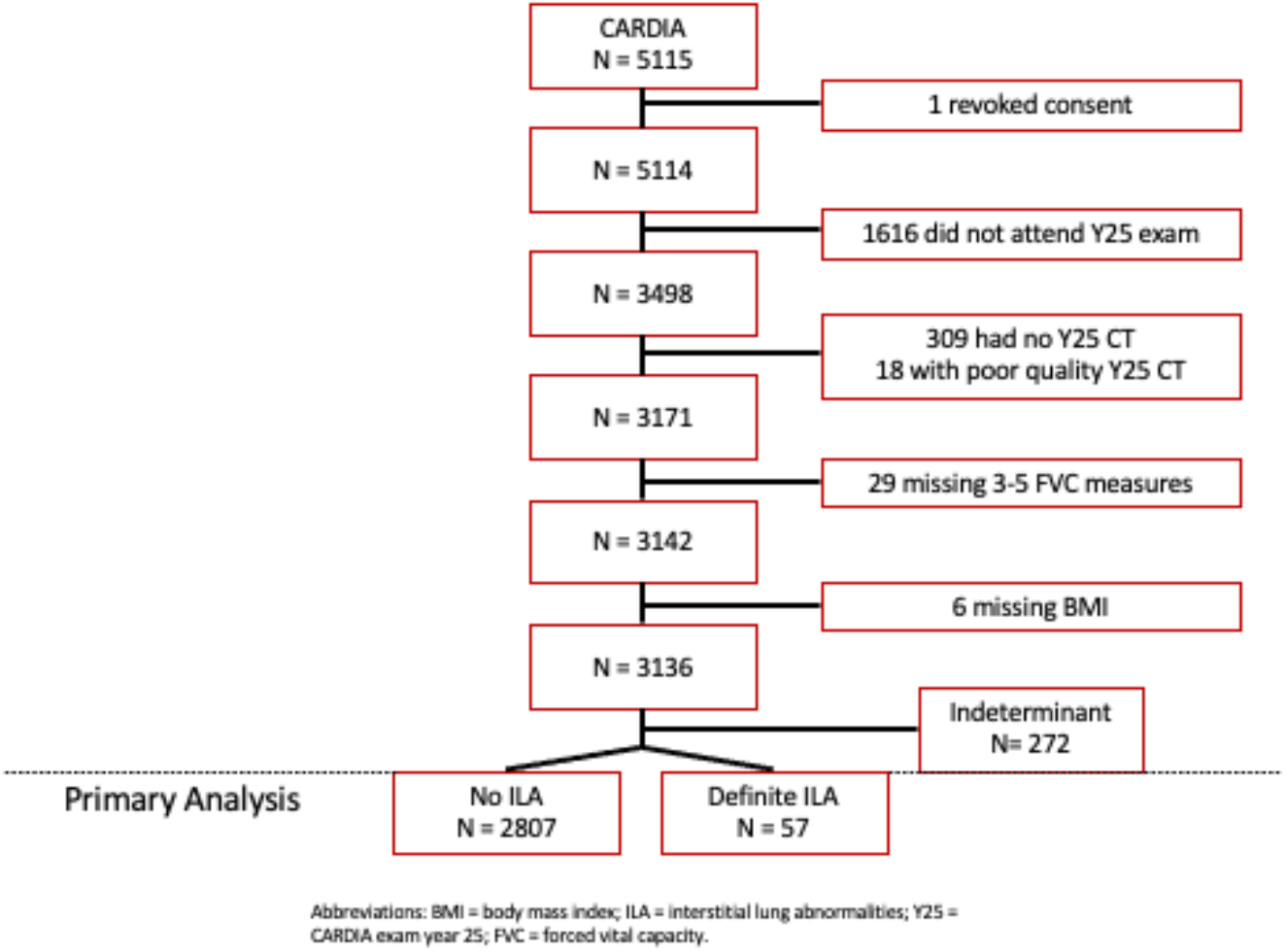
Flow diagram of participant inclusion for the primary analysis. Of the 5,115 participants originally enrolled in the CARDIA study, one revoked consent, 1,616 did not attend the Year 25 (Y25) examination, 309 lacked Y25 chest CT imaging, 18 had poor-quality CT scans, 29 were missing ≥2 forced vital capacity (FVC) measurements, and 6 were missing body mass index (BMI) data. The final analytic cohort included 3,136 participants, classified based on CT findings as having no interstitial lung abnormalities (ILA; n = 2,807), indeterminate ILA (n = 272), or definite ILA (n = 57).

**Figure 2.**
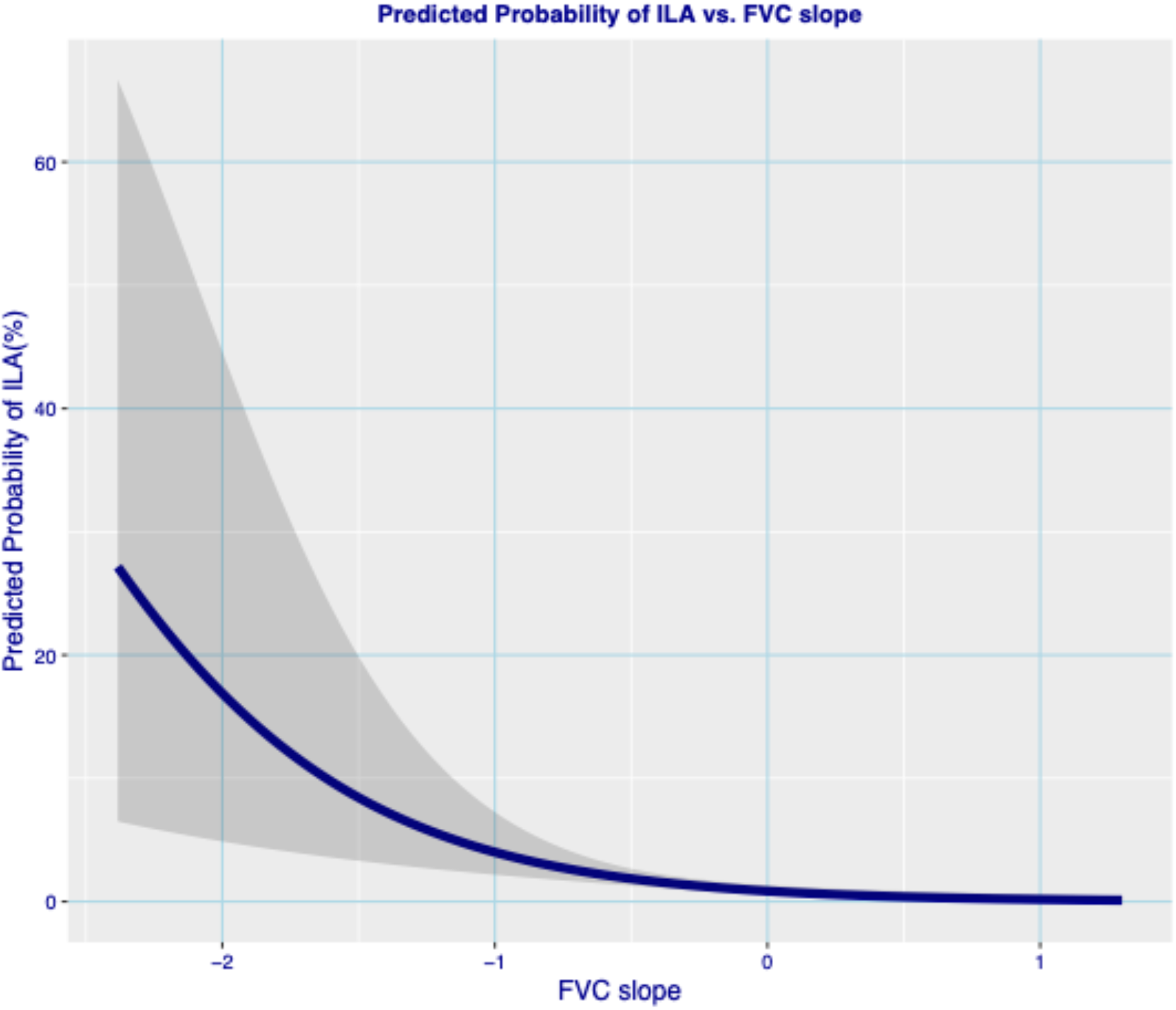
Predicted probability plots of ILA vs. slope FVC. Plot of predicted probability and 95% confidence limits from multivariable logistic regression models predicting definite Year 25 ILA for FVC slope. Covariate values were set at the mean values in the sample.

## DISCUSSION

In this longitudinal cohort study, we report that greater life course decline in lung function was associated with prevalent visually assessed ILA by midlife. These findings expand prior evidence associating impaired lung function with subclinical interstitial changes. In a cohort that is composed of generally younger participants than previously analyzed cohorts, we demonstrate that antecedent physiologic impairment spanning two decades precedes radiologic parenchymal abnormalities previously associated with poor future outcomes.^1–5^

By focusing on visually detected ILA rather than quantitative density measures—which are more prevalent but rely on machine learning-derived density metrics—our study adds clinically-relevant context to earlier analyses.^9^ While QIA may reflect early nonspecific lung density changes, ILA represents a smaller but clinically meaningful subset with established links to mortality and progression to ILD. This distinction underscores the clinical relevance of our findings and highlights the potential of physiologic decline as a precursor to radiologically and clinically significant lung disease. Our results suggest that individuals exhibiting steeper ppFVC decline from early adulthood into midlife may warrant closer monitoring for ILA even before overt symptoms emerge. These data reinforce the concept that subclinical physiologic trajectories can portend later radiologic disease, and they parallel prior work linking impaired lung function trajectories in early life to future risk of emphysema.^9–10^ Previous work showed that ILA can develop and progress over time and are associated with lung function decline,^10^ supporting our findings that long-term physiologic decline precedes clinically meaningful ILA when assessed across an even longer life-course trajectory.

The mechanisms predisposing certain individuals to poor lung health before ILA development remain unclear. Genetic susceptibility, environmental exposures, and/or gene-environment interactions likely contribute to these trajectories. This possibility is underscored by our observation that participants with ILA had a lower proportion of White individuals compared with those without ILA. Elucidating these pathways could clarify why some individuals exhibit early physiologic decline while others maintain stable lung function despite similar exposures.

The CARDIA Lung Study offers a rare life course perspective by integrating repeated measures of lung function, biomarkers, and imaging across decades in a racially diverse population. However, our study still has its limitations. We excluded participants without adequate ppFVC or CT data and those with indeterminate ILA findings, which may have affected our prevalence estimates and introduced selection bias. Additionally, although we adjusted for multiple covariates, residual confounding cannot be excluded.

Future studies should investigate the mechanistic pathways linking early physiologic decline to fibrotic changes and evaluate whether interventions targeting lung function preservation or inflammation modulation can reduce progression of ILA to ILD. Increased frequency of imaging or biomarker collection may help clarify the temporal relationships among lung function decline, inflammation, and radiologic changes. Ultimately, integrating physiologic and biomarker trajectories may improve early detection and inform preventative strategies to reduce ILD risk.

## Abbreviations

ILA: interstitial lung abnormality
CT: computed tomography
ILD: interstitial lung disease
FVC: forced vital capacity
pp: percent predicted

## Notation of Prior Abstract Publication/Presentation

This work was presented as an oral abstract at the American Thoracic Society 2025 International Conference in San Francisco, CA, on 05/20/25. Citation is: Grudzinski K, Liu GY-H, Colangelo L, Selvan KC, Thyagarajan B, Jacobs Jr D, Alexandria S, Putman RK, Hunninghake GM, San Jose Estepar R, Washko GR, Kalhan R, Esposito AJ. Adult life course trajectories of lung function and the development of interstitial lung abnormalities: the CARDIA Lung Study [abstract]. Am J Respir Crit Care Med. 2025;211:A1001.

## Data Availability Statement

All data produced in the present study are available upon reasonable request to the authors and appropriate submission of a dataset request and notice of intent to analyze to CARDIA for approval.

## REFERENCES

1. Podolanczuk AJ, Hunninghake GM, Wilson KC, et al. Approach to the Evaluation and Management of Interstitial Lung Abnormalities: An Official American Thoracic Society Clinical Statement. Am J Respir Crit Care Med. 2025 Jul;211(7):1132–1155.

2. Doyle TJ, Washko GR, Fernandez IE, et al.; COPDGene Investigators. Interstitial lung abnormalities and reduced exercise capacity. Am J Respir Crit Care Med 2012;185:756–762

3. Washko GR, Hunninghake GM, Fernandez IE, et al.; COPDGene Investigators. Lung volumes and emphysema in smokers with interstitial lung abnormalities. N Engl J Med 2011;364:897–906

4. Putman RK, Hatabu H, Araki T, et al. Association Between Interstitial Lung Abnormalities and All-Cause Mortality. JAMA. 2016;315(7):672–681. doi:10.1001/jama.2016.0518

5. Putman RK, Gudmundsson G, Axelsson GT, et al. Imaging Patterns Are Associated with Interstitial Lung Abnormality Progression and Mortality. Am J Respir Crit Care Med. 2019 Jul 15;200(2):175–183. doi: 10.1164/rccm.201809-1652OC

6. Lange P, Celli BR, Agustí García-Navarro À, et al. Lung-function trajectories leading to chronic obstructive pulmonary disease. N Engl J Med. 2015;373:111–122. doi: 10.1056/NEJMoa1411532

7. Kalhan R, Arynchyn A, Colangelo LA, et al. Lung function in young adults predicts airflow obstruction 20 years later. Am J Med. 2010;123(5):468.e1–468.e7. doi:10.1016/j.amjmed.2009.07.037

8. Washko GR, Colangelo LA, San Jose Estepar R, et al. Adult life-course trajectories of lung function and the development of emphysema: The CARDIA Lung Study. Am J Med. 2020;133(2):222-230.e11. doi:10.1016/j.amjmed.2019.06.049

9. Liu GY, Colangelo LA, San Jose Estepar R, et al. Low-normal FVC trajectories starting in early adulthood and risk of future interstitial abnormalities. Am J Respir Crit Care Med. 2023 Oct 1;208(7):816–818. doi: 10.1164/rccm.202304-0771LE.

10. Araki T, Putman RK, Hatabu H, et al. Development and Progression of Interstitial Lung Abnormalities in the Framingham Heart Study. Am J Respir Crit Care Med. 2016 Dec 15;194(12):1514–1522. doi: 10.1164/rccm.201512-2523OC. PMID: 27314401; PMCID: PMC5215030.

11. Friedman GD, Cutter GR, Donahue RP, et al. CARDIA: study design, recruitment, and some characteristics of the examined subjects. J Clin Epidemiol 1988;41:1105–1116. doi: 10.1016/0895-4356(88)90080-7

12. Washko GR, Lynch DA, Matsuoka S, et al. Identification of Early Interstitial Lung Disease in Smokers from the COPDGene Study. Acad Radiol. 2009. doi: 10.1016/j.acra.2009.07.016

13. Podolanczuk AJ, Hunninghake GM, Wilson KC, et al. Approach to the Evaluation and Management of Interstitial Lung Abnormalities: An Official American Thoracic Society Clinical Statement. Am J Respir Crit Care Med. 2025 Jul;211(7):1132–1155. doi: 10.1164/rccm.202505-1054ST

